# Data Farming to Table: Combined Use of a Learning Health System Infrastructure, Statistical Profiling, and Artificial Intelligence for Automating Toxicity and 3-year Survival for Quantified Predictive Feature Discovery from Real-World Data for Patients Having Head and Neck Cancers

**DOI:** 10.1101/2023.10.24.23297349

**Authors:** Charles S Mayo, Shiqin Su, Benjamin Rosen, Elizabeth Covington, Zheng Zhang, Theodore Lawrence, Randi Kudner, Clifton Fuller, Kristy K Brock, Jennifer Shah, Michelle M Mierzwa

## Abstract

**Introduction:** Clinicians iteratively adjust treatment approaches to improve outcomes but to date, automatable approaches for continuous learning of risk factors as these adjustments are made are lacking. We combined a large-scale comprehensive real-world Learning Health System infrastructure (LHSI), with automated statistical profiling, visualization, and artificial intelligence (AI) approach to test evidence-based discovery of clinical factors for three use cases: dysphagia, xerostomia, and 3-year survival for head and neck cancer patients. Our hypothesis was that the combination would enable automated discovery of prognostic features generating testable insights.

**Methods:** Records for 964 patients treated at a single instiution for head and neck cancers with conventional fractionation between 2017 and 2022 were used. Combined information on demographics, diagnosis and staging, social determinants of health measures, chemotherapy, radiation therapy dose volume histogram curves, and treatment details, laboratory values, and outcomes from the LHSI to winnow evidence for 485 candidate prognostic features. Univariate statistical profiling using benchmark resampling to detail confidence intervals for thresholds and metrics: area under the curve (AUC), sensitivity (SN), specificity (SP), F_1_, diagnostic odds ratio (DOR), p values for Wilcoxon Rank Sum (WRS), Kolmogorov-Smirnov (KS), and logistic fits of distributions detailed predictive evidence of individual features. Statistical profiling was used to benchmark, parsimonious XGBoost models were constructed with 10-fold cross validation using training (70%), validation (10%), and test (20%) sets. Probabilistic models utilizing statistical profiling logistic fits of distributions were used to benchmark XGBoost models.

**Results:** Automated standardized analysis identified novel features and clinical thresholds. Validity of automated findings were affirmed with supporting literature benchmarks. Average incidence of dysphagia ≥grade 3 within 1 year of treatment was low (11%). Xerostomia ≥ grade 2 (39% to 16%) and survival ≤ 3 years decreased (25% to 15%) over the time range. Standard planning constraints used limited contribution of those features:: Musc_Constrict_S: Mean[Gy] < 50, Glnd_Submand_High: Mean[Gy] ≤ 30, Glnd_Submand_Low: Mean[Gy] ≤ 10, Parotid_High: Mean[Gy] ≤ 24, Parotid_Low: Mean[Gy] ≤ 10 Additional prognostic features identified for dysphagia included Glnd_Submand_High:D1%[Gy] ≥ 71.1, Glnd_Submand_Low:D4%[Gy] ≥ 55.1, Musc_Constric_S:D10%[Gy] ≥ 56.5, GTV_Low:Mean[Gy] ≥ 71.3. Strongest grade 2 xerostomia feature was Glnd_Submand_Low: D15%[Gy] ≥ 45.2 with a logistic model quantifying a gradual rather than an abrupt increase in probability 13.5 + 0.18 (x-41.0 Gy). Strongest prognostic factors for lower likelihood of death by 3 years were GTV_High: Volume[cc] ≤ 21.1, GTV_Low: Volume[cc] ≤ 57.5, Baseline Neutrophil-Lymphocyte Ratio (NLR) ≤ 5.6, Monocyte-Lymphocyte Ratio (MLR) ≤0.56, Platelet-Lymphocyte ratio (PLR) ≤ 202.5. All predictors had WRS and KS p values < 0.02. Statistical profiling enabled detailing gains of XGBoost models with respect to individual features. Time period reductions in distribution of GTV volumes correlated with reductions in death by 3 years.

**Discussion:** Confirming our hypothesis, automated, standardized statistical profiling of a set of statistical metrics and visualizations supported detailing predictive strength and confidence intervals of individual features, benchmarking of subsequent AI models, and clinical assessment. Association of high dose values to submandibular gland volumes, highlighted relevance as surrogate measures for proximal un-contoured muscles including digastric muscles. Higher values of PLR, NLR, and MLR were associated with lower survival rates. Combined use of Learning Health System Infrastructure, Statistical Profiling and Artificial Intelligence provided a basis for faster, more efficient evidence-based continuous learning of risk factors and development of clinical trial testable hypothesis. Benchmarking AI models with simple probabilistic models provided a means of understanding when results are driven by general areas of overall risk vs. more complex interactions.

## Introduction

Automating learning from large-scale real-world data entered into electronic health record (EHR) systems as part of routine practice is highly desirable to enable rapid and ongoing evidence-driven discovery of features associated with patient outcomes. The development of EHR learning automations will support practice quality improvement and monitoring. In addition, automated evidence-driven discovery could be used to improve hypothesis generation and clinical trial design. Taken together, these developments promise to have a transformative impact on patient care.

In previous work we described the construction of our learning health system (LHS) infrastructure named the Michigan Radiation Oncology Analytics Resource (MROAR) which aggregates, integrates, and harmonizes (AIH) data from the EHR, Radiation Oncology Information System (ROIS), the radiation therapy treatment planning system (TPS) and other systems. The goal was to enable electronic learning from large scale, comprehensive real-world data sets. We implemented standardizations as part of clinical workflows to reduce noise and missingness in manually entered data. This marked approaching AIH scalability systematically as a “data farming”, rather than a more ad-hoc “data mining”, operation^1^.

In a subsequent study, we coupled that system to an algorithm that combined statistical profiling with artificial intelligence to create an explainable AI approach to detail prognostic factors of patients being treated for head and neck cancers having emergency department visits during or within 90 days of the end of treatment. ^2^

We expanded upon the methods, algorithms, and visualizations from our prior work to improve detection and evaluation of candidate prognostic factors. By integrating statistical profiling, including standardized visualizations and fittings, along with AI approaches layered on top of the comprehensive LHS, our overarching aim is to build a standardized, clinically interpretable approach to drive evidence-based discovery and hypothesis generation from large-scale, multi-factor “real-world” data sets. ^3^

We tested the ability of the approach to highlight previously under-recognized prognostic features in our local data in three use cases: 1) dysphagia, 2) xerostomia, and 3) three-year survival rates for patients treated for head and neck cancers. We tested features identified for supporting evidence in the literature, expanding the range of locally monitored features. Statistical profiling was used to quantify confidence intervals and actionable thresholds. Benchmarking of AI models with respect to mechanistic statistical models was used to quantify gains in AI models and to gauge impact of missing data.

In this work we demonstrate we show that coupling the standardized numerical and visualization methodology to the comprehensive, large-scale learning health system provides insights to better guide treatment planning and monitoring throughout patient care. Clinical incorporation of standardizations in data entry, aggregation, analysis and reporting, creates a data “farm to table” approach for automating clinical discovery from real-world data. In addition, the use of standardized, scalable, automated approaches for continuous evidence-based learning could support ongoing practice quality monitoring with positive implications for accreditation, Medicare payment programs, and other quality improvement activities.

## Methods

Records for 964 patients treated for head and neck cancers with conventional fractionation between 2017 and 2022 were used in the study operating under a hospital IRB. Patient characteristics are summarized in Table 1. Patients were predominantly male (74%) and white (91%).

**Table 1.** Patient characteristics.

| Patients |  | 974 |
| --- | --- | --- |
| Sex | Female | 252 (26%) |
|  | Male | 722 (74%) |
| Stage | I | 91 ( 9%) |
|  | II | 101 (10%) |
|  | III | 206 (21%) |
|  | IV | 469 (48%) |
|  | Metastatic | 107 (11%) |
| Race | White | 884 (91%) |
|  | Black | 30 ( 3%) |
|  | Asian | 18 ( 2%) |
|  | Unspecified | 42 ( 4%) |
| Ethnicity | Hispanic | 17 ( 2%) |
|  | Non-Hispanic | 945 (97%) |
|  | Unspecified | 12 ( 1%) |

We examined 468 prognostic features spanning 9 domains (Table 2) to identify predictors for dysphagia ≥ grade 3, xerostomia ≥ grade 2, and death within 3 years of treatment. TG-263 nomenclature standardizations were used for structures and DVH metrics.^4^ Applying the Operational Ontology for Oncology (O3), DVH curves were represented as an absolute volume and a set of Dx%[Gy] values for a set of standardized percentage values (100%,99.5%, 99-96% in 1% increments, 95%-5% in 5% increments, 4%-1% in 1% increments, 0.5% and 0%). D0%[Gy] corresponds to Max[Gy] and D100%[Gy] corresponds to Min[Gy]. ^5^ Bilateral structures (parotid and submandibular glands) were categorized as _High vs. _Low according to their relative median dose. Following TG-263 nomenclature, for plans with multiple GTV and PTV volumes treated to different dose levels, the volumes receiving the highest and lowest dose were categorized as xxx_High and _Low and if three volumes, then xxx_Mid were also used. Single target volumes were categorized as _High.

**Table 2:** Comprehensive per patient feature set.

| Domain | Features |
| --- | --- |
| Demographics | Sex, Race, Ethnicity, Martial Status, Insurance, Age |
| Location | Postal Code, Distance to hospital |
| Disease Characteristics | Diagnosis and Staging, Volume of GTVs, CTVs, PTVs, number of GTVs |
| Substances | Smoking history, smoking max pack/day, alcohol history, max oz/week of alcohol |
| Body Habitus | Value nearest course start and end for: Height, weight, BMI |
| Chemotherapy Agents | Carboplatin, Cisplatin, Paclitaxel, Bevacizumab, Cyclophosphamide, Docetaxel, Fluorouracil, Gemcitabine, Methotrexate, Nivolumab, Pembrolizumab, Vincristine |
| Laboratory Values (course start and end) | Value nearest course start and at 2 months following end of treatment for: Neutrophile (Absolute and Percent), Lymphocytes (Absolute and Percent), Platelets, Hematocrit, Hemoglobin, White blood cell count, Red blood cell count, Mean corpuscular volume, Monocytes Absolute, Eosinophils Absolute, Basophiles (Absolute and Percent), Albumin, Chloride, Potassium, Sodium, Creatinine, Blood urea nitrogen, Total Bilirubin, Neutrophil-Lymphocyte ratio, Platelet-Lymphocyte ratio, Monocyte-Lymphocyte ratio, Albumin-Bilirubin (ALBI) |
| Treatment | Number of re-simulations, Median dose to GTV, CTV and PTV |
| O3 Formatted Organ at Risk DVHs curves | Superior constrictor, inferior constrictor, parotid glands, submandibular glands, oral cavity, larynx, esophagus, brainstem |

All calculations were carried out in Python 3.6, with statistical profiling was carried out for each feature. Profiles had three components: 1) bootstrap resampling of thresholds and derivative statistics detailing evidence and confidence intervals were tabulated for filtering and sorting, 2) visualization of distributions and logistic fits supporting clinical evaluation, and 3) visualizations box-whisker plots by year to highlight evolving practice changes.

Endpoints analyzed included overall survival and toxicity endpoints of dysphagia and xerostomia according to physician-graded toxicity prospectively scored at each clinical visit including consults, on-treatment visits, and follow-up visits.

For component 1, a set of statistical measures was calculated for each of the 1000 samples of each feature. In each sample, the Youden index for a receiver operator characteristic (ROC) curve was used to identify an optimal threshold. Incidence (I), area under the curve (AUC), sensitivity (SN), specificity (SP), positive predictive value (PPV), negative predictive value (NPV), F_1_, Matthew’s Correlation Coefficient^6–9^, Diagnostic Odds Ratio (DOR)^10,11^, p values for Wilcoxon Rank Sum Test (WRS), and Kolmogorov-Smirnov (KS) tests were calculated for each sample. In addition, we calculated the relative predictive difference (RPD) with respect to the overall incidence for each sample as

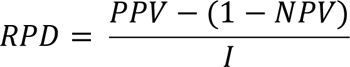

to gauge the ability of the threshold to consistently categorize the feature value region of greatest incidence.

Mean, standard deviation, median, and 25% and 75% quantiles were calculated from distribution of metric values over all samples. The set of bootstrapped sampled statistical metrics and confidence intervals were used to evaluate univariate evidence for predictive strength of each feature.

For component 2, distributions for those with and without the outcome were compared in a histogram for each feature. Median threshold from component 1 (*x*_0_) was plotted for reference. The distribution with respect to the feature value (x) was fitted (*P*_*max*_, *dx*) to an increasing (*P*_I_ (*x*)) and a decreasing (*P*_D_ (*x*)) logistic curve.

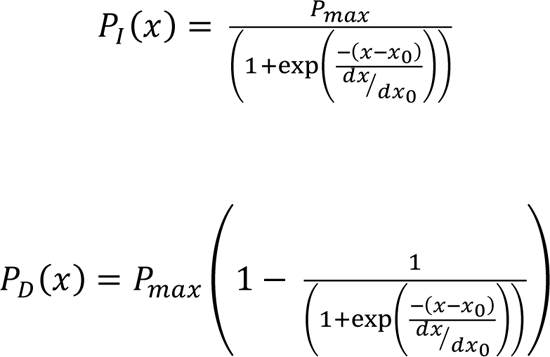

where,

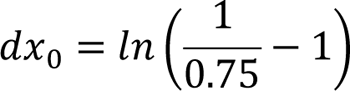

Definition of *dx*_0_ means *P*_I_ (*x*_0_ + *dx*_0_) = 0.75 ∗ *P*_*max*_. The curve fit selected to represent the distribution (P) was set equal to *P*_I_ or *P*_D_ according to whether the median PPV or (1-NPV) was larger. P was plotted along with the histogram. The plots were used to visualize the strength of evidence for each feature.

SN and an SP prioritized feature sets were created from component 1 tabulations, filtering for median (AUC ≥ 0.6, KS p≤ 0.05, and WRS p ≤0.05), and SN ≥ 0.6 or SP ≥ 0.6 respectively. Features passing both filters were included according to whether SN or SP was larger. Features were displayed in order of importance using SN*DOR*P_max_ or SP*DOR*P_max_ respectively. For structures in each set, only the DVH metric that was most important was retained. For laboratory values, only the most important value at the start of the course or two months following the end of the course was retained for any given test.

We defined a feature clinical relevance (FCR) score (table 3) to reflect concordance with literature and clinician experience.

**Table 3:** Feature Clinical Relevance (FCR) scoring.

| Feature Clinical Relevance Value | Description |
| --- | --- |
| **1.25** | Feature supported by literature and not currently incorporated in local clinical practice or outcomes modeling. |
| **1.00** | Feature supported by literature |
| **0.50** | Feature not supported by literature but judged by clinicians as likely relevant. |
| **-1.00** | Feature not supported by literature. |
| **-1.50** | Feature not supported by literature and judged by clinicians as likely not relevant. |

Baseline XGBoost models were constructed and compared in the context of statistical profiling of individual features for each SN and SP prioritized feature set individually and for the combination of SP and SN feature sets. Models were also constructed to compare results for data sets where missing values were imputed with median values to sets limited to the subset of records not requiring imputation.

Twenty-fold cross-validation with training and testing sets (80%,20%) was carried out with each XGBoost model. Hyperparameters were tuned to reduce overfitting. Since incidence is low, Synthetic Minority Oversampling Technique (SMOTE) of training sets was used to mitigate under-sampling in model creation. Statistics were aggregated across the folds for AUC, SN, and SP for testing sets. XGBoost reported importance of the features in the training set was aggregated across the folds.

Significance of improvement in AUC, SN, and SP in models over univariate statistical profiling and of XGBoost over the more statistically driven logistic fit model was evaluated with a student t-test (p<0.05).

## Results

Incidence of dysphagia, xerostomia, and Kaplan-Meier 3-year overall survival by year of course start is shown in Figure 1 along with the makeup of patient cohorts by stage. Staging system changed from AJCC7 to AJCC8 in 2020. Incidence of grade 3 dysphagia requiring a feeding tube was ∼11% over the time range, while xerostomia and survival ≤3 years decreased.

**Figure 1.**
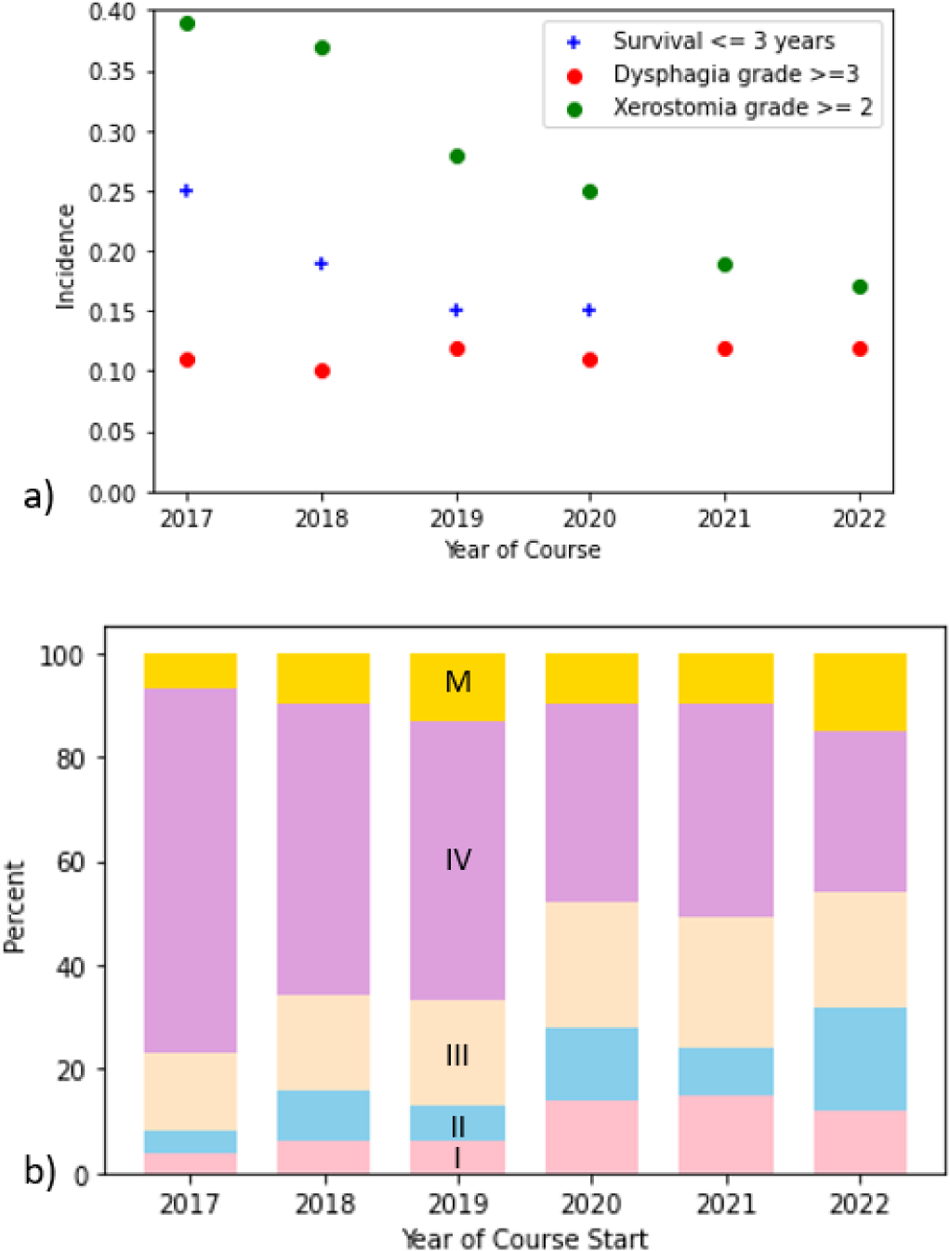
Change by Year of Course start of a) Incidence of Dysphagia grade ≥ 3 (red circle), Xerostomia grade ≥ 2 (green circle) and Survival ≤ 3 years (blue plus sign) compared to b) mix of cancer stage I (pink), II (blue), III(beige), IV(lavender), Documented Distant Metastasis (M, yellow)

Figure 2 illustrates the combined numerical and visualization approach for one of the features evaluated for predicting dysphagia, Ipsilateral Submandibular Gland: D1%[Gy]. Augmenting statistical metrics with visualizations improved communication with stakeholders and evaluation of clinical relevance and interpretation of numerically identified features. Thresholds supported clinical evaluation in the context of clinician experience providing a basis for evaluating applicability in clinical use or in a trial design. DOR and RPD highlighted features with stronger contrasts in incidence on either side of identified thresholds. Evaluating standardized DVH metrics along the whole DVH curve enabled selecting the specific regions with strongest evidence. Use of a logistic model plotted alongside distributions provided ready means for visualizing strength of evidence for a feature with respect to the median threshold. Comparing changes in incidence over time (Figure 1) with changes in distributions of feature values (Figure 2c) supported explain-ability of time dependence.

**Figure 2.**
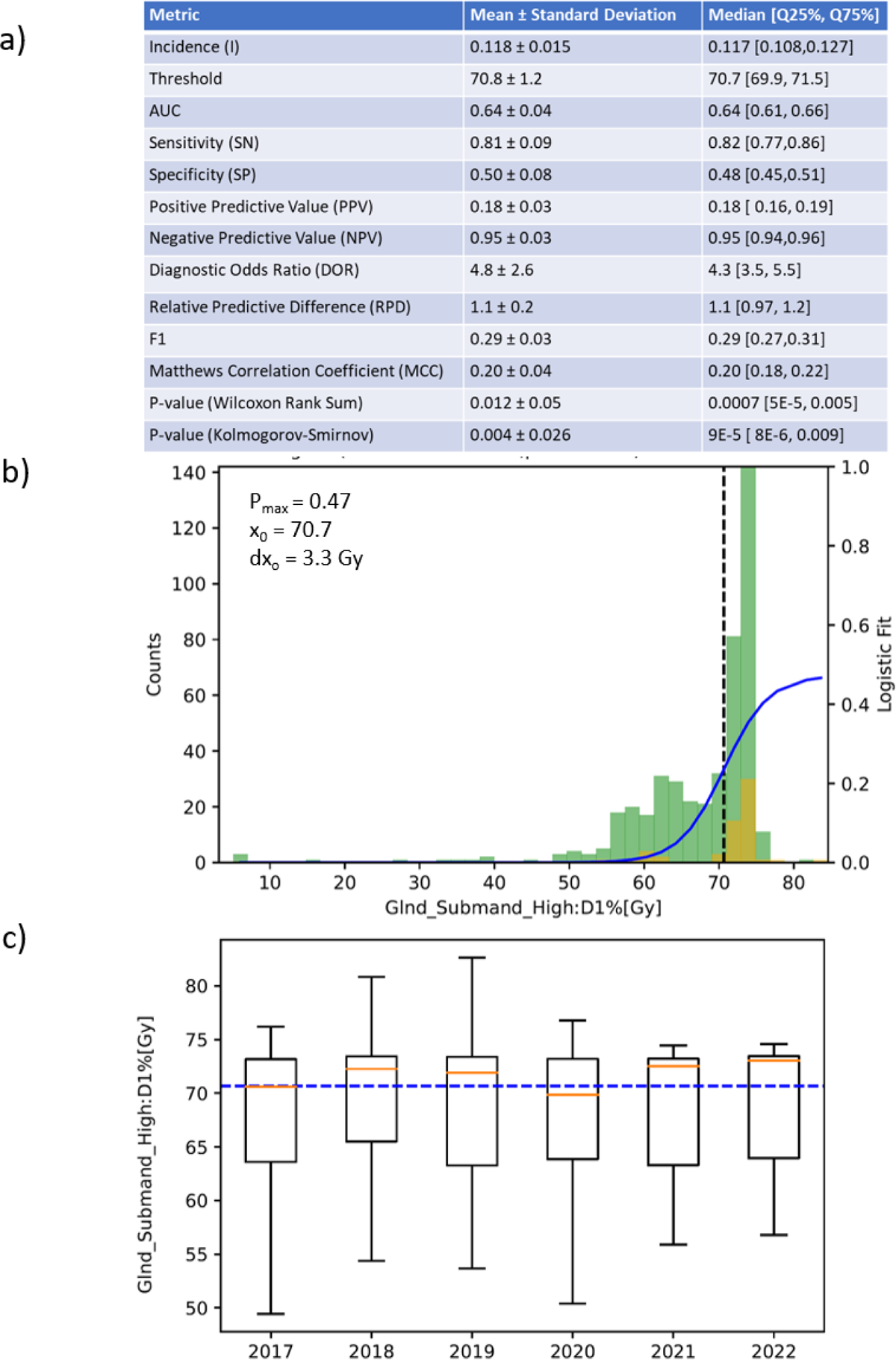
Statistical profiling results for one of the features evaluated for predicting grade 3 dysphagia. Combining (a) tabulations of boot strap resampling of a common set of thresholds and statistical characterization metrics, with (b) visualizations of distributions and logistic model fit (solid blue line) for those with (yellow) and without (green) toxicity and (c) box-whisker plots of distributions by year with respect to median threshold (dashed blue line) supported clinical evaluation of features and explain-ability of subsequently developed models.

Standard planning constraints limit reduced likelihood of a dataset containing high dose values for several metrics. Current constraints include Larynx: Mean[Gy] <20 (3), Musc_Constrict_S: Mean[Gy] < 50, Musc_Constrict_S: D25%[Gy] < 50, Oral_Cavity: Mean[Gy] ≤ 30Gy, Glnd_Submand_High: Mean[Gy] ≤ 30, Glnd_Submand_Low: Mean[Gy] ≤ 10, Parotid_High: Mean[Gy] ≤ 24, Parotid_Low: Mean[Gy] ≤ 10, Esophagus: Mean[Gy] ≤ 20. Glnd_Submand_Low and Parotid_Low limits were added after 2017. Table 4 summarizes statistical profiling and modeling results for SN and SP prioritized features. Visualizations of distributions and logistic fits for all features are shown in the appendix.

**Table 4:** Standardized comparison of AI algorithm results in the context of statistical profiling of features for a) Dysphagia ≥ grade 3, b) Xerostomia ≥ grade 2, and c) Survival ≤3 Years with number of patients in the labeled subset in parenthesis. Missingness is detailed for each feature as well as fraction meeting threshold limit. Bootstrap resampling of Thresholds, AUC, SN, SP, and DOR highlight strength of evidence for each with confidence intervals as median [25% quantile, 75% quantile]. Gradual vs step changes in logistic model fits are highlighted for dx0 greater or less than x0 respectively. Feature clinical relevance (FCR) was scored to reflect concordance with literature and clinician experience.

| a) Dysphagia ≥ grade 3 (819) |  | SN Prioritized Set |  |  |  |  |  |  |
| --- | --- | --- | --- | --- | --- | --- | --- | --- |
| Feature | Fraction With Feature | More likely for rel to x0 (Fraction meeting limit) | AUC | SN | SP | DOR | $dx_0/x_0$ | FCR |
| GlnD_Submand_High: D1%[Gy] | 0.62 | ≥ 70.7 [69.9,71.6] (0.57) | 0.64 [0.61,0.66] | 0.82 [0.77,0.86] | 0.48 [0.45,0.51] | 4.3[3.5,5.5] | 0.05 | 1.25 |
| GlnD_Submand_Low: D4%[Gy] | 0.84 | ≥ 55.1 [ 51.5, 56.4] (0.52) | 0.70 [0.68,0.72] | 0.86 [0.78,0.91] | 0.51 [0.44,0.60] | 6.1[5.0,8.1] | 0.13 | 1.25 |
| Larynx: Max[Gy] * | 0.88 | ≥ 65.6 [ 64.3, 67.7] (0.34) | 0.67 [0.65,0.69] | 0.63 [0.55,0.69] | 0.63 [0.55,0.69] | 4.1[3.5,4.9] | 0.11 | 1 |
| Parotid_Low: Max[Gy] | >0.99 | ≥ 58.2 [ 57.5, 59.3] (0.6) | 0.66 [0.64, 0.67] | 0.84 [0.77,0.88] | 0.53 [0.36,0.66] | 4.2[3.4,5.2] | 0.63 | 1 |
| Musc_Constrict_S: D50%[Gy] | >0.99 | ≥ 47.4 [ 41.0, 48.7] (0.52) | 0.67 [0.66,0.69] | 0.79 [0.71,0.89] | 0.51 [0.36,0.66] | 4.2[3.5,5.5] | 0.30 | 1 |
| Oral_Cavity: D30%[Gy] | 0.98 | ≥ 37.1 [28.1,41.2] (0.49) | 0.63 [0.61,0.65] | 0.72 [0.62,0.91] | 0.55 [0.30, 0.85] | 3.6[2.8,5.2] | 0.50 | 1.25 |
| Days to Max Toxicity * | 1.0 | ≥ 29 [29, 32] (0.41) | 0.72 [0.70,0.73] | 0.78 [0.74, 0.86] | 0.64 [0.61,0.65] | 6.1[5.3,7.7] | 9.4 | 1 |
| PTV_High: Mean[Gy] * | 1.0 | ≥ 71.2 [68.8,71.3] (0.42) | 0.60 [0.58,0.62] | 0.63 [0.58,0.67] | 0.60 [0.59,0.62] | 2.6[2.2,3.0] | < 0.01 | 1 |
| XGB_Baseline (53% Imputed) |  |  | 0.79 [0.77 0.83] | 0.84 [0.74 0.89] | 0.69 [0.61 0.79] | 11.3 [7.7,16.4] |  |  |
| XGB_Baseline Not imputed Subset |  |  | 0.79 [0.76 0.82] | 0.81 [0.73 0.89] | 0.70 [0.58 0.80] | 11.4 [8.2, 18.0] |  |  |

|  |  | SP Prioritized Set |  |  |  |  |  |  |
| --- | --- | --- | --- | --- | --- | --- | --- | --- |
| Feature | Fraction With Feature | Less likely for rel to x0 (Fraction meeting limit) | AUC | SN | SP | DOR | $dx_0/x_0$ | FCR |
| GlnD_Submand_High: D30%[Gy] | 0.62 | ≤ 70.4 [67.8,70.8] (0.74) | 0.68 [0.65,0.70] | 0.56 [ 0.49, 0.70] | 0.79 [0.61,0.82] | 4.3[3.5,5.2] | 0.08 | 1.25 |
| GlnD_Submand_Low: D90%[Gy] | 0.84 | ≤ 34.3 [24.4, 50.0] (0.74) | 0.65 [0.63,0.67] | 0.53 [0.43,0.62] | 0.76 [0.67,0.84] | 3.8[3.1,4.7] | 0.54 | 1.25 |
| Musc_Constrict_S: Max[Gy] | >0.99 | ≤ 21.8 [14.9,24.7] (<0.01) | 0.61 [0.59,0.63] | 0.50 [0.43,0.64] | 0.77 [0.59,0.82] | 3.1[2.7,3.6] | 0.10 | 0.5 |
| Chemo Includes Paclitaxel | 1.0 | No (0.75) | 0.61 [0.59,0.62] | 0.43 [0.40,0.47] | 0.78 [0.77,0.79] | 2.7[2.3,3.1] | NA | 1 |
| Oral_Cavity: D5%[Gy] | 0.98 | ≤ 63.0 [63.0,63.2] (0.38) | 0.65 [0.63,0.67] | 0.62 [0.58, 0.66] | 0.66 [0.64,0.68] | 3.1[2.7,3.6] | 0.17 | 1 |
| Chemo Includes CarboPlatin | 1.0 | No (0.6) | 0.60 [0.58,0.62] | 0.58 [0.55,0.61] | 0.63 [0.62,0.64] | 1.7[1.5,2.0] | NA | 1 |
| Parotid_Low: D45%[Gy] | >0.99 | ≤ 19.9 [16.5, 20.0] (0.6) | 0.63 [0.61,0.65] | 0.66 [0.60, 0.79] | 0.62 [0.46,0.64] | 3.0[2.6,3.6] | 0.50 | 1.0 |
| XGB_Baseline (53% Imputed) |  |  | 0.67 [0.62 0.71] | 0.74 [0.62 0.89] | 0.61 [0.45 0.72] | 5.1 [3.5 7.0] |  |  |
| XGB_Baseline Not imputed Subset |  |  | 0.67 [0.63 0.71] | 0.74 [0.58 0.84] | 0.62 [0.48 0.77] | 4.8 [3.6 7.1] |  |  |

| Combined SN and SP sets |  | AUC | SN | SP | DOR |
| --- | --- | --- | --- | --- | --- |
| XGB_Baseline (53% Imputed) |  | 0.79 [0.77 0.83] | 0.79 [0.71 0.89] | 0.74 [0.66 0.81] | 11.6 [7.8 15.93] |
| XGB_Baseline Not imputed Subset |  | 0.80 [0.77 0.85] | 0.81 [0.70 0.91] | 0.78 [0.65 0.88] | 13.4 [7.5, 22.5] |

| b) Xerostomia ≥ grade 2 (890) |  | SN Prioritized Set |  |  |  |  |  |  |
| --- | --- | --- | --- | --- | --- | --- | --- | --- |
| Feature | Fraction With Feature | More likely for rel to x0 (Fraction meeting limit) | AUC | SN | SP | DOR | $dx_0/x_0$ | FCR |
| Days to Max Toxicity | 1.0 | ≥ 23 [19,26] (0.50) | 0.71 [0.70,0.72] | 0.87 [0.82,0.90] | 0.52 [0.49,0.58] | 6.3[5.5,7.4] | 0.22 | 1 |
| GlnD_Submand_Low: D25%[Gy] | 0.84 | ≥ 41.0 [40.0,44.4] (0.57) | 0.60 [0.59, 0.62] | 0.69 [0.60, 0.74] | 0.5 [0.46,0.58] | 2.3[2.0,2.6] | 0.89 | 1.25 |
| Baseline XGB (Impute 26%) |  |  | 0.75 [0.73 0.77] | 0.86 [0.77 0.91] | 0.58 [0.53 0.68] | 8.64[6.6,12.0] |  |  |
| XGB_Baseline Not imputed Subset |  |  | 0.75 [0.73 0.77] | 0.87 [0.80 0.91] | 0.58 [0.51 0.66] | 9.6 [7.3 11.5] |  |  |

|  |  | SP prioritized set |  |  |  |  |  |  |
| --- | --- | --- | --- | --- | --- | --- | --- | --- |
| Feature | Fraction Missing | Less likely for rel to x0 (Fraction meeting limit) | AUC | SN | SP | DOR | $dx_0/x_0$ | FCR |
| GlnD_Submand_Low:D75%[Gy] | 0.84 | ≤ 28.9 [ 21.2, 30.7] (0.62) | 0.61 [0.60,0.63] | 0.55 [0.51,0.72] | 0.67 [0.49,0.70] | 2.5[2.2,2.8] | 1.1 | 1.25 |
| Baseline XGB (Impute 26%) |  |  | 0.60 [0.57 0.62] | 0.49 [0.41 0.82] | 0.71 [0.32 0.78] | 2.6 [2.2 3.1] |  |  |
| XGB_Baseline Not imputed Subset |  |  | 0.59 [0.56 0.61] | 0.51 [0.42 0.80] | 0.71 [0.35 0.79] | 2.48 [2.1,3.2] |  |  |

| Combined SN and SP sets |  | AUC | SN | SP | DOR |
| --- | --- | --- | --- | --- | --- |
| Baseline XGB (Impute 26%) |  | 0.76 [0.73 0.78] | 0.84 [0.77 0.91] | 0.60 [0.53 0.66] | 8.4 [6.4,12.0] |
| XGB_Baseline Not imputed Subset |  | 0.75 [0.73 0.77] | 0.86 [0.77 0.91] | 0.60 [0.56 0.68] | 8.9 [7.0 11.7] |

| c) Survival ≤ 3 years (519) |  | SN prioritized set |  |  |  |  |  |  |
| --- | --- | --- | --- | --- | --- | --- | --- | --- |
| Feature | Fraction With Feature | More likely for (Fraction meeting limit) | AUC | SN | SP | DOR | $dx_0/x_0$ | FCR |
| Platelet-Lymphocyte Ratio at Course start * | 0.89 | ≥ 202.5 [197.4, 212.1] (0.42) | 0.64 [0.62,0.66] | 0.66 [ 0.6,0.72] | 0.64 [0.59,0.67] | 3.3[2.8,4.0] | 0.36 | 1.25 |
| XGB_Baseline (Imputed 11%) |  |  | 0.61 [0.56 0.65] | 0.63 [0.5 0.83] | 0.68 [0.35 0.73] | 2.85 [2.14 4.30] |  |  |
| XGB_Baseline Not imputed Subset |  |  | 0.60 [0.56 0.64] | 0.63 [0.50 0.83] | 0.65 [0.39 0.72] | 2.83 [2.19 4.14] |  |  |

|  |  | SP prioritized set |  |  |  |  |  |  |
| --- | --- | --- | --- | --- | --- | --- | --- | --- |
| Feature | Fraction With Feature | Less likely for rel to x0 (Fraction meeting limit) | AUC | SN | SP | DOR | $dx_0/x_0$ | FCR |
| GTV_High:Volume[cc] | 0.47 | ≤ 21.1 [16.9,28.8] (0.78) | 0.65 [0.61,0.69] | 0.52 [ 0.43,0.60] | 0.84 [0.77, 0.92] | 6.3[4.5,9.4] | 2.5 | 1.25 |
| Neutrophile at Course start | 0.89 | ≤ 72.8[63.4, 74.3] (0.77) | 0.61 [0.59,0.64] | 0.49 [0.38, 0.81] | 0.79 [0.42, 0.86] | 3.7[3.0,4.6] | 0.42 | 1.25 |
| Neutrophile-Lymphocyte Ratio at Course Start | 0.89 | ≤ 5.6 [4.6, 5.7] (0.81) | 0.63 [0.60, 0.65] | 0.44 [ 0.9,0.52] | 0.84 [ 0.78,0.87] | 4.1[3.3,4.9] | 5.6 | 1.25 |
| Monocyte-Lymphocyte Ratio at course start | 0.89 | ≤ 0.55 [0.53, 0.57] (0.63) | 0.61 [0.58, 0.63] | 0.57[0.52, 0.63] | 0.67 [0.63,0.70] | 2.6[2.2,3.1] | 0.93 | 1.25 |
| XGB_Baseline (Imputed 57%) |  |  | 0.62 [0.56 0.65] | 0.54 [0.44 0.78] | 0.76 [0.46 0.86] | 3.8 [2.8, 5.0] |  |  |
| XGB_Baseline Not imputed Subset |  |  | 0.68 [0.62 0.74] | 0.57 [0.43 0.71] | 0.84 [0.79 0.89] | 8.8 [5.7, 15.6] |  |  |

Table 4: Standardized comparison of AI algorithm results in the context of statistical profiling of features for a) Dysphagia ≥ grade 3, b) Xerostomia ≥ grade 2, and c) Survival ≤3 Years with number of patients in the labeled subset in parenthesis.
| Combined SN and SP sets |  | AUC | SN | SP | DOR |
| --- | --- | --- | --- | --- | --- |
| XGB_Baseline (Imputed 57%) |  | 0.62 [0.57 0.66] | 0.56 [0.44 0.78] | 0.74 [0.46 0.85] | 4.0 [2.7,5.5] |
| XGB_Baseline Not imputed Subset |  | 0.71 [0.65 0.76] | 0.71 [0.57 0.71] | 0.82 [0.76 0.89] | 9.4 [5.9, 15.6] |

### Dysphagia

High dose to submandibular glands was not among the features monitored in standard planning constraints but was found to be significant (p<< 0.01). Glnd_Submand_High: D1%[Gy] and D30%[Gy] values equal to 71 Gy had SN and SP of 0.82 and 0.79, respectively. While current constraints focus on Larynx: Mean[Gy], Max[Gy] ≥ 65.5 was also found to be a predictor (p<<0.01). Thresholds in Oral_Cavity dose D30%[Gy] ≥ 37 (SN = 0.72, p<< 0.01) and D5%[Gy] ≤ 63 (SP = 0.66, p<<0.01) indicated further reductions beyond standard constraints could be beneficial.

Patients not treated with carboplatin or cisplatin were less likely (p<<0.01) to have dysphagia with median SP equal to 0.63[0.62,0.64] and 0.81[0.80,0.82] respectively. Cisplatin was used infrequently.

Risk was greater for maximum toxicity occurring late in the course or after completion of treatment (≥ 29 days), consistent with known clinical practice.

Including SP-prioritized features did not significantly improve XGBoost models compared to SN-prioritized features alone. Use of imputation did not significantly change model predictions.

Having more than one GTV (p< 0.04), weight change (>-7.3[-7.3,-6.3] kg, p< 0.01), BMI change (>-2.1[-2.1,-1.5], p<0.01), and weight (< 71.6[66.6, 76.6] kg, p< 0.02) were significant but did not have AUC, SN, SP > 0.6. Sex, race, ethnicity, and insurance status did not convey greater risk (p>0.3). Observed age threshold was 62.3 [60.4, 65.6] but was not significant (p>0.09). Stage was not a significant (p>0.38) predictor.

### Xerostomia

In the context of dose distributions driven by current planning constraints, low doses to submandibular glands had the strongest associations with xerostomia for Glnd_Submand_Low: D25%[Gy] ≥ 41 (SN = 0.69, p < 0.01) and Glnd_Submand_Low: D75%[Gy] ≤ 28.9 (SP=0.67, p< 0.01). Parotid_High: D50%[Gy] ≤ 24.3[22.4,24.9] was marginally significant (WRS p = 0.07, KS p = 0.04) and had SN = 0.78[0.57, 0.80], but AUC = 0.58[0.57,0.60]. Parotid_Low: D50% ≤ 12.0[11.5,16.7] Gy was significant (p< 0.01) with SN = 0.78[0.57, 0.80], but AUC= 0.58[0.57,0.60].

Figure 2 (a,b) illustrates that distribution differences in Glnd_Submand_Low: D25%[Gy] were more pronounced than D50% consistent with routine use of the Mean[Gy] constraint in routine planning. Logistic fits illustrated that the percent risk for xerostomia due to submandibular gland dose increased gradually (*d*_*x*0_⁄_*x*0_ = 0.89) rather than as a step. Risk of xerostomia associated with Glnd_Submand_Low: D25%[Gy] increased gradually as 13.5% + 0.18 (x-41.0 Gy)%.

Observed reduction in xerostomia by year (figure 1a), correlated with reduction in the lower quartile of doses to parotid and submandibular glands decreased over time (2 c-f). Reductions for for submandibular glands were most pronounced.

Stage was not a significant (p>0.5) predictor. Risk was greater for maximum toxicity occurring late in the course (≥ 23 days after start of treatment).

### Survival ≤ 3 Years

Patients with GTV_Low and GTV_High volumes ≤ 57.5 cc and ≤21.1 cc respectively were less likely to have survival ≤ 3 years (SP = 0.84, p<0.02 and 0.78, p<0.001 respectively). Figure 3 a-d highlights that relative to the thresholds, more patients with larger GTV volumes presented in 2017 than in subsequent years.

**Figure 3.**
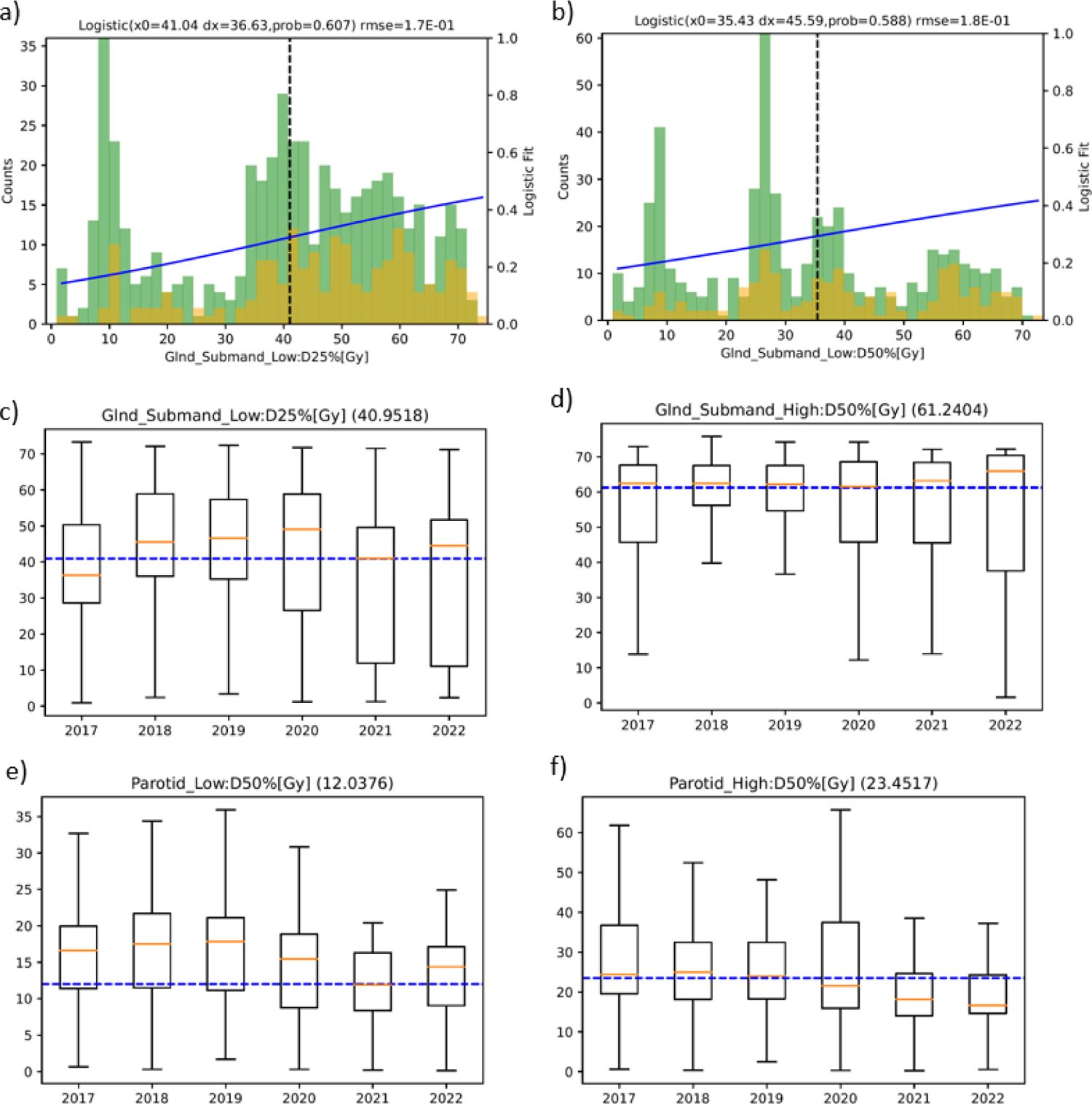
Use of statistical profiling visualizations supporting practice changes over time of features contributing to xerostomia

**Figure 4.**
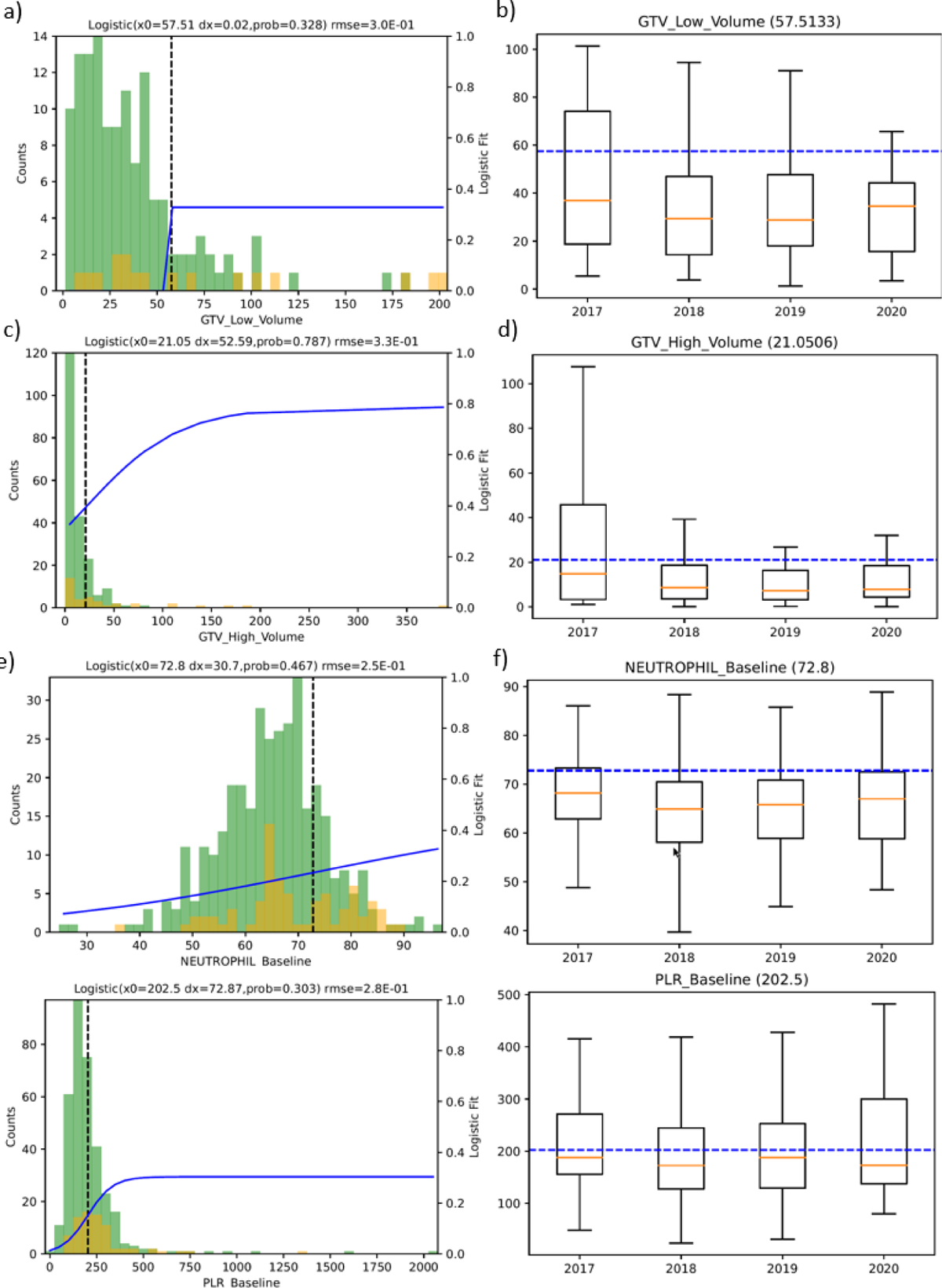
Features predicting Survival ≤3 year

Significant improvement in ML models for non-imputed vs. imputed sets highlighted the importance of GTV volumes to models.

Stage IV was marginally significant with (WRS p< 0.03, KS p > 0.08) and AUC= 0.57[0.55,0.59], SN= 0.59[0.55,0.62], SP=0.56[0.54,0.57]. The average fraction of patients with Stage IV disease 2017-2018 was 55% 2017-2018, decreasing to 31% in 2020-2022.

We had not previously included laboratory values among the markers monitored for anticipating outcomes. Patients with platelet lymphocyte ratios (PLR) ≥ 202.5 or neutrophile counts ≥ 4.8 at the course start were more likely (SN = 0.66, p< 0.01 and SN = 0.65, p<0.01) to have survival ≤ 3 years. Similarly, patients with neutrophile-lymphocyte ratio (NLR) ≤ 5.6, percent neutrophile ≤ 72.8, or monocyte-lymphocyte ratios (MLR) ≤ 0.55 were less likely to have survival ≤ 3 years with SP = 0.84, p<0.01 and 0.79, p<0.03 and 0.67, p<0.04 respectively. Patients with baseline platelet ≤ 278[262,302] were less likely to have survival ≤ 3 years (SP = 0.70[0.62,0.78],).

## Discussion

The combined use of the standardized LHSI + Statistical Profiling + AI approach demonstrated the ability to automate hypothesis-generating clinical insights from comprehensive, large-scale multi-feature real-world data sets. Successfully benchmarking automated findings with similar findings in the literature (FCR≥1.0) supports efficacy of the method.

### Dysphagia

Significance (p<<0.01) of high doses to the submandibular glands and the contralateral parotid gland for dysphagia was a novel finding from our experience. Threshold dose levels were much higher than associated with xerostomia, and hypothesis generating. Dysphagia is complex, and patients often describe dry mouth as contributory to their dysphagia, but these higher doses may also be consistent with muscle or nerve toxicities. Submandibular glands are adjacent to the digastric, geniohyoid, stylohyoid, and mylohyoid muscles. These are not routinely contoured but have been implicated in the swallowing mechanism. This leads to the hypothesis that high doses to the submandibular glands may act as surrogates for dose to the adjacent musculature. If true then constraining high doses to submandibular glands may be used to reduce dysphagia incidence without needing to increase the number of musculature structures contoured.

A few publications support this novel hypothesis (FCR=1.25). In a study of 300 patients with oropharyngeal cancer, with delineation of several swallowing muscle groups, Dale et al found that genioglossus V35Gy[%], anterior digastric muscle V60Gy[%], middle constrictor V49Gy[%], and superior constrictor muscle V70Gy[%] were associated with increased risk but did not provide confidence interval quantified thresholds.^12^ They did not differentiate between doses to components of bilateral muscle groups. They found a continuous risk model with two inputs V69Gy[%] for mylo/geniohyoid complex and age out had an AUC of 0.835, performing better than a binary model for mylo/geniohyoid complex V69Gy[%] >79.5 and age of > 62 years. Values and confidence intervals for SN and SP for models were not provided.

In a study of 90 patients Hedström et al. found that constructing a model using mean doses to the contralateral parotid and supraglottic larynx, and maximum dose to the contralateral anterior digastric muscle had an AUC = 0.64-0.67.^13^ Mean[Gy] ≥60 for submandibular glands and Mean[Gy] ≥40 for digastric muscles were the most significant predictors. Mean[Gy] pharyngeal constrictor muscle, the larynx, the supraglottic larynx, the epiglottis, and Max[Gy] to the contralateral submandibular gland predicted moderate and severe dysphagia by VFS (AUC = 0.71-0.80).

Examining other features, in a study of 31 patients Schwarz et al. reported adding Oral Cavity: V30Gy[%] < 65 and V35Gy[%] < 35 as well as superior constrictor V55Gy[%] < 80 and V65Gy[%] < 30 as planning constraints based on their study to reduce the incidence of dysphagia. They did not quantify risks with SN or SP. ^14^

### Xerostomia

Parotid sparing is widely adopted as the primary means of limiting incidence of xerostomia. Given dose constraints targeting parotids, additional gains are evident with reductions to submandibular glands.

Longitudinal decreases on the lower quartile of submandibular gland doses correlated with observed reductions in the incidence of high-grade xerostomia. This reflected practice changes over the same time period.

In a study of 114 patients with 76 receiving contralateral submandibular gland sparing to Mean[Gy] ≤ 30.7, Gensheimer found significantly reduced incidence.^15^ Kim et al. reported results from a randomized study of 236 patients having elective nodal irradiation of IB nodes.^16^ They found that lower Mean[Gy] for ipsilateral (51.5 vs 67.9) and contralateral (45.2 vs. 55.7) were associated with the incidence of grade 2 xerostomia (9.9 vs. 29.2%)

Our results are suggestive that differential dosing to subvolumes of contralateral submandibular glands D25%[Gy] < 41 and D75%[Gy] < 28.9 may be beneficial (FCR=1.25).

### Survival

Stage IV was a predictor for survival ≤3 years, but more specifically GTV_Low and GTV_High volumes greater than identified thresholds were associated with shorter survival. Significance of missing values in ML models is suggestive of the potential value of recording GTV volumes before surgical resection to include in models. Data was suggestive that elevated platelets, neutrophils, and monocytes were associated with shorter survival but were less important than GTV volumes. PTV and CTV volumes did not emerge as predictive indicating that the underlying disease rather than the volume of irradiated tissue may be more predictive. Literature supported identified values that are not currently incorporated into our local models (LCR = 1.25)

We have previously shown that the median GTV volume of the primary tumor is a significant predictor of locoregional failure (LRF) (Mierzwa, CCR, 2022). In another retrospective analysis of 137 patients with nasopharyngeal cancer, Laconelli et al. identified GTV volume < 43.2 cc as a prognostic factor for local control at 5 years. ^17^ Romesser et al. examined the prognostic utility of GTV volumes for primary (GTV-P) and nodal (GTV-N) disease to augment TNM classification. ^18^ They found that GTV-P < 32.9 cc was a significant predictor for overall survival as well as for local control, but not volume of GTV-N. Studer et al. examined the potential of a GTV volume-based system of staging for predicting overall survival in a cohort of 172 patients treated for Head and Neck Cancers. They demonstrated significant differences in Kaplan Meier survival curves categorizing primary and total GTV volumes into cohorts of 1-15 cc, 15-70cc and > 70cc.^19^ Our findings that GTV_High (i.e. primary GTV) < 21.2 cc and GTV_Low (i.e. total GTV) < 57.5 cc support their hypothesis that incorporation of GTV volumes into prognostic models could improve survival models based on TNM staging alone (FCR=1.25).

In a study of 433 patients with oropharyngeal cancer, Shoultz-Henly et al. found that freedom from distant metastasis and overall survival was decreased for patients with pre-treatment platelet > 350 x 10^9^/L ^20^. Haddad found in a study of 46 patients that 2-year overall survival was 89% vs. 61% for patients with NLR < 5. ^21^ Bardash et al. carried out a meta-analysis of 13 studies from 4541 patients and reported that elevated PLR was associated with poorer survival. Median PLR threshold from the studies ranged from 11 to 186.2 with a median of 146.2. ^22^ A meta-analysis showed that larger PLR, NLR, and MLR values were associated with lower overall survival. ^23^ A study of 476 patients by Yu et al. demonstrated that a lymphocyte-monocyte ratio < 3.8 (corresponding to MLR > 0.26) was associated with lower overall survival. ^24^ Our findings supported prognostic value of PLR,NLR, and MLR and quantified thresholds for clinical guidance (FCR=1.25)

## Summary

There are significant challenges for the application of AI with real-world data. Real-world data may be sparse (not all patients have all features or labels). Further, AI algorithms may not provide improved insights beyond statistical analysis for all data sets.^25,26^ We found that detailing inputs ahead of the application of AI algorithms in decision frameworks enabled a more objective assessment of results and insights on surrogacy of features.

Use of only a single metric (e.g. AUC) and the neglect of methods to quantify confidence intervals undermines the ability to objectively assess evidence and likely reproducibility of findings. We recommend that at minimum SN, SP, DOR, WRS and KS with confidence intervals determined with bootstrap resampling be used to augment AUC as a reporting metric. Optimally the complete set of metrics used here, including thresholds and confidence intervals, would be included in reporting.

Ability to automate statistical and AI discovery is only as strong as the consistent entry of feature values. When clinical standardized methods for entry are not used or are endlessly personalized, then everyone is harmed by forcing manual rather than automated aggregation of data. This example focused on comprehensive feature sets where data was reliably entered showing the potential of automated approaches.

Use of the feature clinical relevance (FCR) was a novel and valuable metric for benchmarking features identified with the automation. For any given feature, the FCR metric is likely to evolve as clinical factors become relevant with clinical practice changes. This points to the value of journals adopting standardized article meta-data approaches (e.g. based on O3) to facilitate searching and scoring that could be incorporated into automation. This would enable developing automated analysis approaches that readily incorporate literature values with local data to improve rapid discovery and validation.

Standardized detailed analysis and visualizations revealed practice patterns changed. For accreditation agencies development of radiation therapy specific practice quality metrics is an area of interest.^27–31^ To optimize participation, metrics need to be both clinically relevant and possible to gather without excessive effort. We believe application standardized LHSI + Statistical Profiling + AI systems across a wider range of facilities could be transformative for both.

Both the Quantitative Analysis of Normal Tissue Effects in the Clinic (QUANTEC) and the Pediatric Normal Tissue Effects in the Clinic (PENTEC) highlighted non-standard reporting of dose delivered to organs at risk and small data sets not representing the overall population as issues in the development of models from aggregated experience.^32–35^ By using the combination of LHSI + statistical profiling + AI to lower the effort to create standardized data sets and reporting methods, such systems could significantly improve the ability to learn from aggregated experience by expanding the range of academic and non-academic clinics able to contribute.

## Data Availability

Data on results produced in the present study are available upon reasonable request to the authors. Access to underlying raw data is dependent on conditions of use and will require completion of institutional data use agreement.

---

Appendix Statistical Profiling Distribution visualizations from SP and SN screened features

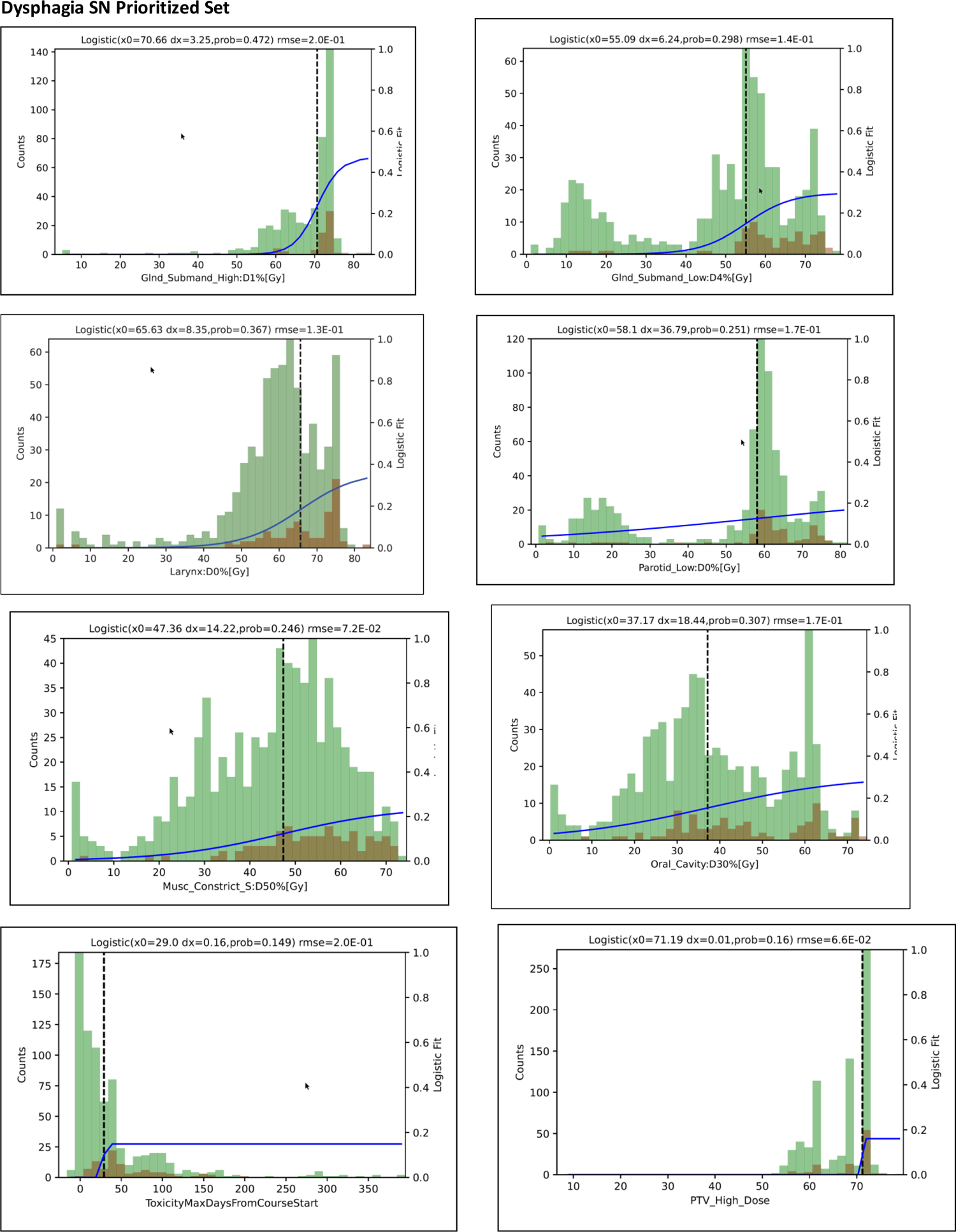

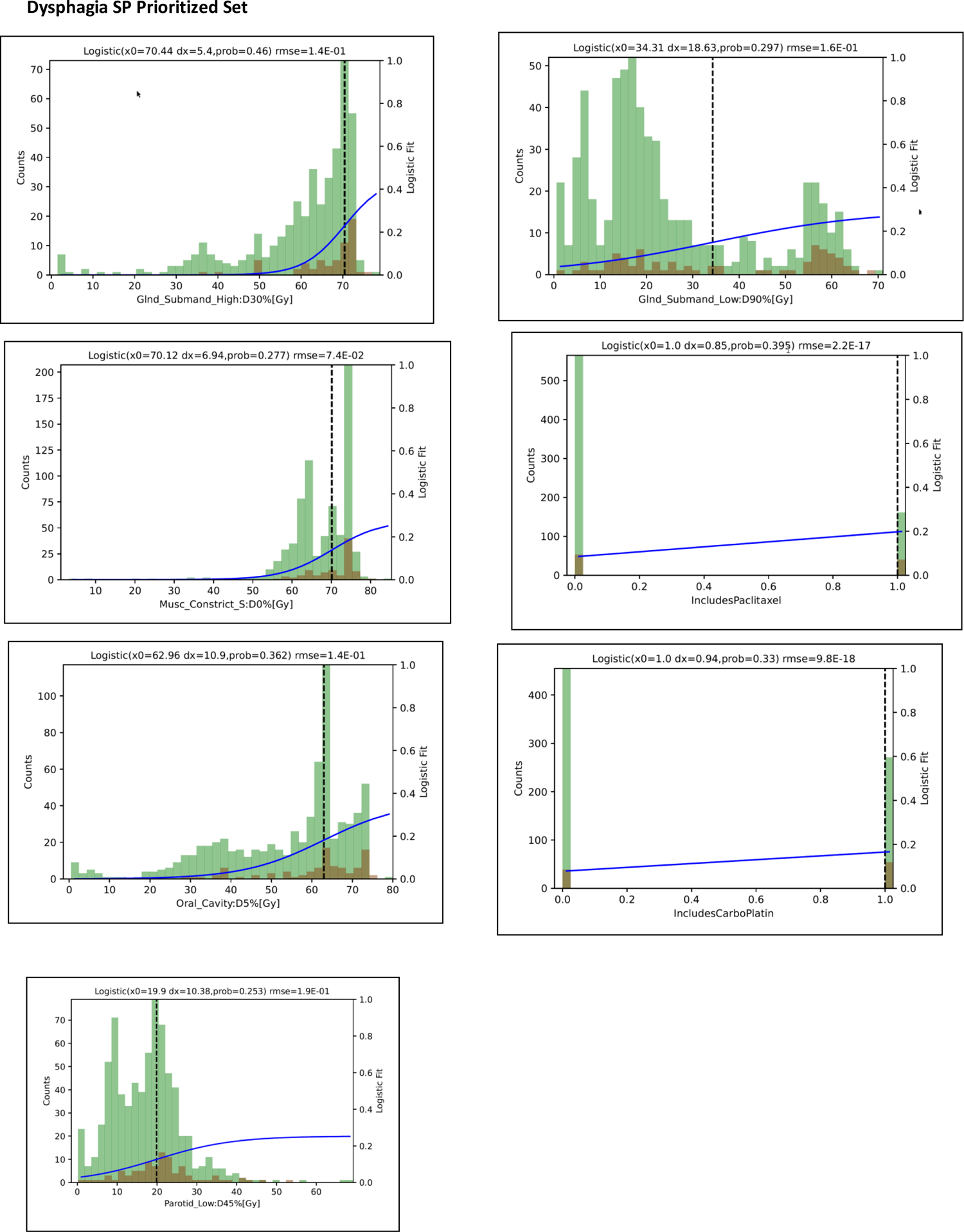

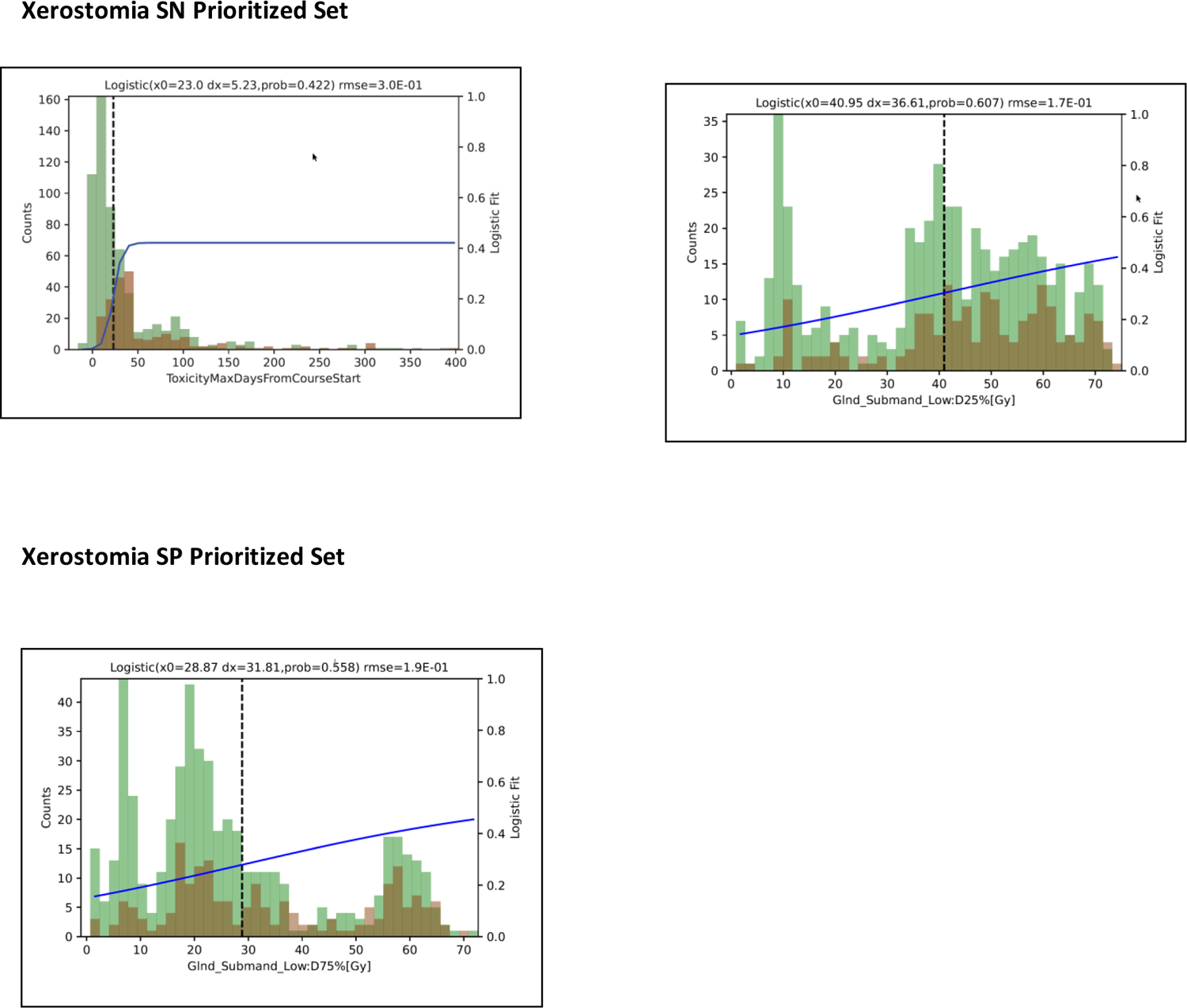

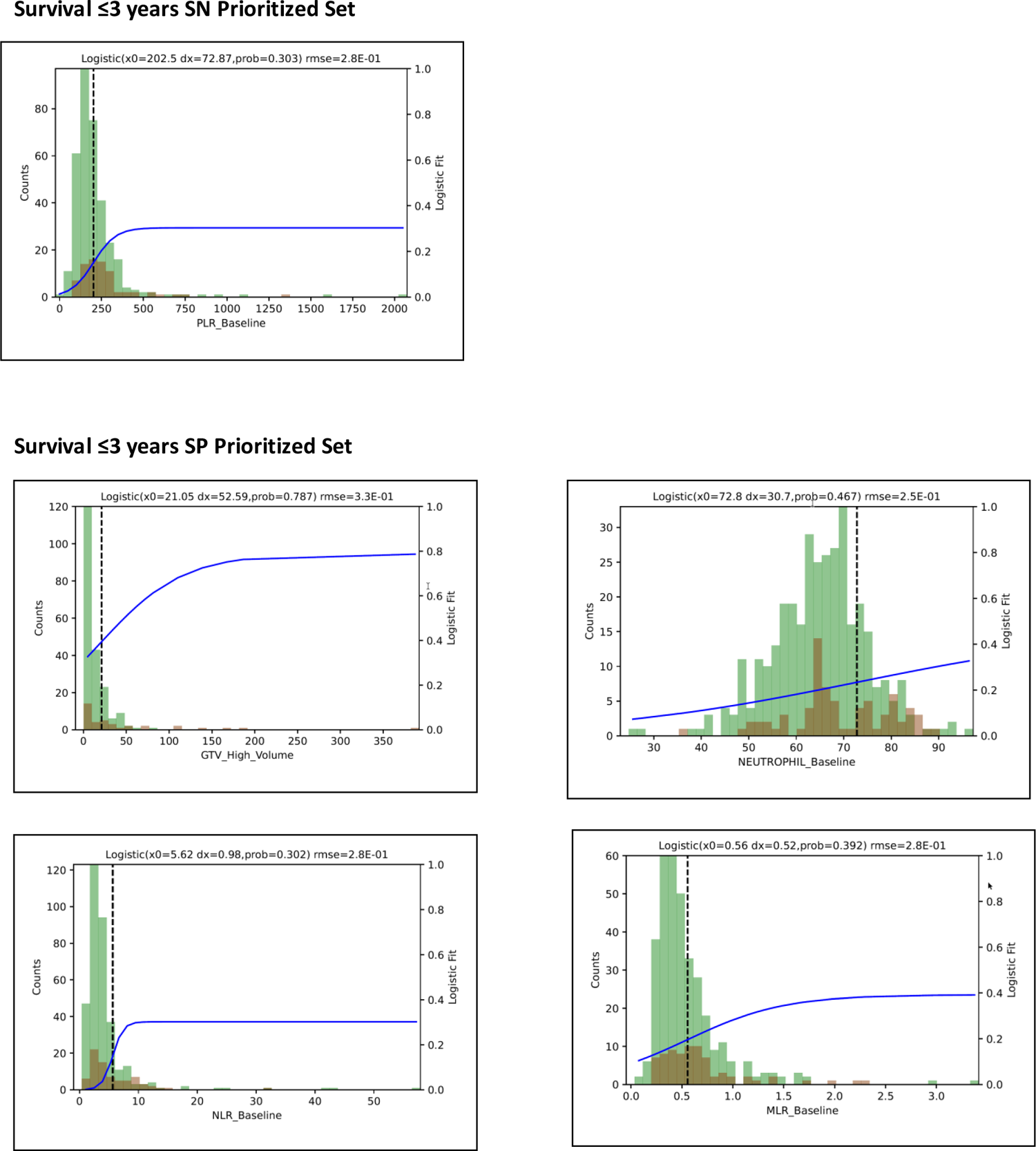

## References

1. Mayo CS, Kessler ML, Eisbruch A, Weyburne G, Feng M, Hayman JA, Jolly S, El Naqa I, Moran JM, Matuszak MM, Anderson CJ, Holevinski LP, McShan DL, Merkel SM, Machnak SL, Lawrence TS, Ten Haken RK. The big data effort in radiation oncology: Data mining or data farming? Adv Radiat Oncol. 2016 Dec;1(4):260–271. PMCID: PMC5514231

2. Mayo CS, Mierzwa M, Yalamanchi P, Evans J, Worden F, Medlin R, Schipper M, Schonewolf C, Shah J, Spector M, Swiecicki P, Mayo K, Casper K. Machine Learning Model of Emergency Department Use for Patients Undergoing Treatment for Head and Neck Cancer Using Comprehensive Multifactor Electronic Health Records. JCO Clin Cancer Inform. Wolters Kluwer; 2023 Jan;(7):e2200037.

3. Mayo CS, Matuszak MM, Schipper MJ, Jolly S, Hayman JA, Ten Haken RK. Big Data in Designing Clinical Trials: Opportunities and Challenges. Front Oncol. 2017;7:187. PMCID: PMC5583160

4. Mayo CS, Moran JM, Bosch W, Xiao Y, McNutt T, Popple R, Michalski J, Feng M, Marks LB, Fuller CD, Yorke E, Palta J, Gabriel PE, Molineu A, Matuszak MM, Covington E, Masi K, Richardson SL, Ritter T, Morgas T, Flampouri S, Santanam L, Moore JA, Purdie TG, Miller RC, Hurkmans C, Adams J, Wu QRJ, Fox CJ, Siochi RA, Brown NL, Verbakel W, Archambault Y, Chmura SJ, Dekker AL, Eagle DG, Fitzgerald TJ, Hong T, Kapoor R, Lansing B, Jolly S, Napolitano ME, Percy J, Rose MS, Siddiqui S, Schadt C, Simon WE, Straube WL, St. James ST, Ulin K, Yom SS, Yock TI. American Association of Physicists in Medicine Task Group 263: Standardizing Nomenclatures in Radiation Oncology. Int J Radiat Oncol Biol Phys. 2018 Mar 15;100(4):1057–1066. PMCID: PMC7437157

5. Mayo CS, Feng MU, Brock KK, Kudner R, Balter P, Buchsbaum JC, Caissie A, Covington E, Daugherty EC, Dekker AL, Fuller CD, Hallstrom AL, Hong DS, Hong JC, Kamran SC, Katsoulakis E, Kildea J, Krauze AV, Kruse JJ, McNutt T, Mierzwa M, Moreno A, Palta JR, Popple R, Purdie TG, Richardson S, Sharp GC, Shiraishi S, Tarbox L, Venkatesan AM, Witztum A, Woods KE, Yao J, Farahani K, Aneja S, Gabriel PE, Hadjiiski L, Ruan D, Siewerdsen JH, Bratt S, Casagni M, Chen S, Christodouleas J, DiDonato A, Hayman J, Kapoor R, Kravitz S, Sebastian S, Von Siebenthal M, Xiao Y. Operational Ontology for Oncology (O3) - A Professional Society Based, Multi-Stakeholder, Consensus Driven Informatics Standard Supporting Clinical and Research use of “Real - World” Data from Patients Treated for Cancer: Operational Ontology for Radiation Oncology. Int J Radiat Oncol Biol Phys. 2023 May 25;S0360–3016(23)00525–4. PMID: 37244628

6. Chicco D, Jurman G. The advantages of the Matthews correlation coefficient (MCC) over F1 score and accuracy in binary classification evaluation. BMC Genomics. 2020 Jan 2;21(1):6. PMCID: PMC6941312

7. Chicco D, Jurman G. The Matthews correlation coefficient (MCC) should replace the ROC AUC as the standard metric for assessing binary classification. BioData Min. 2023 Feb 17;16(1):4. PMCID: PMC9938573

8. Chicco D, Tötsch N, Jurman G. The Matthews correlation coefficient (MCC) is more reliable than balanced accuracy, bookmaker informedness, and markedness in two-class confusion matrix evaluation. BioData Min. 2021 Feb 4;14(1):13. PMCID: PMC7863449

9. Boughorbel S, Jarray F, El-Anbari M. Optimal classifier for imbalanced data using Matthews Correlation Coefficient metric. PloS One. 2017;12(6):e0177678. PMCID: PMC5456046

10. Glas AS, Lijmer JG, Prins MH, Bonsel GJ, Bossuyt PMM. The diagnostic odds ratio: a single indicator of test performance. J Clin Epidemiol. 2003 Nov;56(11):1129–1135. PMID: 14615004

11. Šimundić AM. Measures of Diagnostic Accuracy: Basic Definitions. EJIFCC. 2009 Jan 20;19(4):203–211. PMCID: PMC4975285

12. MD Anderson Head and Neck Cancer Symptom Working Group. Beyond mean pharyngeal constrictor dose for beam path toxicity in non-target swallowing muscles: Dose-volume correlates of chronic radiation-associated dysphagia (RAD) after oropharyngeal intensity modulated radiotherapy. Radiother Oncol J Eur Soc Ther Radiol Oncol. 2016 Feb;118(2):304–314. PMCID: PMC4794433

13. Hedström J, Tuomi L, Finizia C, Olsson C. Identifying organs at risk for radiation-induced late dysphagia in head and neck cancer patients. Clin Transl Radiat Oncol. 2019 Nov;19:87–95. PMCID: PMC6804434

14. Schwartz DL, Hutcheson K, Barringer D, Tucker SL, Kies M, Holsinger FC, Ang KK, Morrison WH, Rosenthal DI, Garden AS, Dong L, Lewin JS. Candidate dosimetric predictors of long-term swallowing dysfunction after oropharyngeal intensity-modulated radiotherapy. Int J Radiat Oncol Biol Phys. 2010 Dec 1;78(5):1356–1365. PMCID: PMC4034521

15. Gensheimer MF, Liao JJ, Garden AS, Laramore GE, Parvathaneni U. Submandibular gland-sparing radiation therapy for locally advanced oropharyngeal squamous cell carcinoma: patterns of failure and xerostomia outcomes. Radiat Oncol Lond Engl. 2014 Nov 26;9:255. PMCID: PMC4262974

16. Kim D, Keam B, Ahn SH, Choi CH, Wu HG. Feasibility and safety of neck level IB-sparing radiotherapy in nasopharyngeal cancer: a long-term single institution analysis. Radiat Oncol J. 2022 Dec;40(4):260–269. PMCID: PMC9830035

17. Iacovelli NA, Cicchetti A, Cavallo A, Alfieri S, Locati L, Ivaldi E, Ingargiola R, Romanello DA, Bossi P, Cavalieri S, Tenconi C, Meroni S, Calareso G, Guzzo M, Piazza C, Licitra L, Pignoli E, Carlo F, Orlandi E. Role of IMRT/VMAT-Based Dose and Volume Parameters in Predicting 5-Year Local Control and Survival in Nasopharyngeal Cancer Patients. Front Oncol. 2020;10:518110. PMCID: PMC7541899

18. Romesser PB, Qureshi MM, Subramaniam RM, Sakai O, Jalisi S, Truong MT. A prognostic volumetric threshold of gross tumor volume in head and neck cancer patients treated with radiotherapy. Am J Clin Oncol. 2014 Apr;37(2):154–161. PMCID: PMC5014357

19. Studer G, Lütolf UM, El-Bassiouni M, Rousson V, Glanzmann C. Volumetric staging (VS) is superior to TNM and AJCC staging in predicting outcome of head and neck cancer treated with IMRT. Acta Oncol Stockh Swed. 2007;46(3):386–394. PMID: 17450476

20. Shoultz-Henley S, Garden AS, Mohamed ASR, Sheu T, Kroll MH, Rosenthal DI, Gunn GB, Hayes AJ, French C, Eichelberger H, Kalpathy-Cramer J, Smith BD, Phan J, Ayoub Z, Lai SY, Pham B, Kies M, Gold KA, Sturgis E, Fuller CD. Prognostic value of pretherapy platelet elevation in oropharyngeal cancer patients treated with chemoradiation. Int J Cancer. 2016 Mar 1;138(5):1290–1297. PMCID: PMC4779600

21. Haddad CR, Guo L, Clarke S, Guminski A, Back M, Eade T. Neutrophil-to-lymphocyte ratio in head and neck cancer. J Med Imaging Radiat Oncol. 2015 Aug;59(4):514–519. PMID: 25908427

22. Bardash Y, Olson C, Herman W, Khaymovich J, Costantino P, Tham T. Platelet-Lymphocyte Ratio as a Predictor of Prognosis in Head and Neck Cancer: A Systematic Review and Meta-Analysis. Oncol Res Treat. 2019;42(12):665–677. PMID: 31550732

23. Kumarasamy C, Tiwary V, Sunil K, Suresh D, Shetty S, Muthukaliannan GK, Baxi S, Jayaraj R. Prognostic Utility of Platelet–Lymphocyte Ratio, Neutrophil–Lymphocyte Ratio and Monocyte–Lymphocyte Ratio in Head and Neck Cancers: A Detailed PRISMA Compliant Systematic Review and Meta-Analysis. Cancers. 2021 Aug 19;13(16):4166. PMCID: PMC8393748

24. Yu B, Ma SJ, Khan M, Gill J, Iovoli A, Fekrmandi F, Farrugia MK, Wooten K, Gupta V, McSpadden R, Kuriakose MA, Markiewicz MR, Al-Afif A, Hicks WL, Seshadri M, Ray AD, Repasky EA, Singh AK. Association of pre-treatment lymphocyte-monocyte ratio with survival outcome in patients with head and neck cancer treated with chemoradiation. BMC Cancer. 2023 Jun 21;23(1):572. PMCID: PMC10286492

25. Kann BH, Hosny A, Aerts HJ. Artificial Intelligence for Clinical Oncology. Cancer Cell. 2021 Jul 12;39(7):916–927. PMCID: PMC8282694

26. Sarker IH. Machine Learning: Algorithms, Real-World Applications and Research Directions. SN Comput Sci. 2021; 2(3):160. PMCID: PMC7983091

27. Hong DS, Boike T, Dawes S, Klash SJ, Kudner R, Okoye C, Rosu-Bubulac M, Watanabe Y, Wright JL, Jennelle RL. Accreditation Program for Excellence (APEx): A Catalyst for Quality Improvement. Pract Radiat Oncol. 2021;11(2):101–107. PMID: 33279669

28. Kapoor R, Moghanaki D, Rexrode S, Monzon B, Ray M, Hulick PR, Albuquerque K, Rosenthal SA, Palta JR, Hagan MP. Quality Improvements of Veterans Health Administration Radiation Oncology Services Through Partnership for Accreditation With the ACR. J Am Coll Radiol JACR. 2018 Dec;15(12):1732– 1737. PMID: 30100162

29. Potters L, Gaspar LE, Kavanagh B, Galvin JM, Hartford AC, Hevezi JM, Kupelian PA, Mohiden N, Samuels MA, Timmerman R, Tripuraneni P, Vlachaki MT, Xing L, Rosenthal SA, American Society for Therapeutic Radiology and Oncology, American College of Radiology. American Society for Therapeutic Radiology and Oncology (ASTRO) and American College of Radiology (ACR) practice guidelines for image-guided radiation therapy (IGRT). Int J Radiat Oncol Biol Phys. 2010 Feb 1;76(2):319–325. PMID: 20117284

30. Hayman JA, Dekker A, Feng M, Keole SR, McNutt TR, Machtay M, Martin NE, Mayo CS, Pawlicki T, Smith BD, Kudner R, Dawes S, Yu JB. Minimum Data Elements for Radiation Oncology: An American Society for Radiation Oncology Consensus Paper. Pract Radiat Oncol. 2019 Nov;9(6):395–401. PMID: 31445187

31. Evans SB, Martin DD, Kudner R. The Standard Prescription and APEx Accreditation: One Hand Washes the Other. Pract Radiat Oncol. 2019 Nov;9(6):389–391. PMID: 31233894

32. Bentzen SM, Constine LS, Deasy JO, Eisbruch A, Jackson A, Marks LB, Haken RKT, Yorke ED. Quantitative Analyses of Normal Tissue Effects in the Clinic (QUANTEC): An Introduction to the Scientific Issues. Int J Radiat Oncol Biol Phys. 2010 Mar 1;76(3 Suppl):S3–S9. PMCID: PMC3431964

33. Constine LS, Ronckers CM, Hua CH, Olch A, Kremer LCM, Jackson A, Bentzen SM. Pediatric Normal Tissue Effects in the Clinic (PENTEC): An International Collaboration to Analyse Normal Tissue Radiation Dose-Volume Response Relationships for Paediatric Cancer Patients. Clin Oncol R Coll Radiol G B. 2019 Mar;31(3):199–207. PMCID: PMC7556704

34. Mayo C, Martel MK, Marks LB, Flickinger J, Nam J, Kirkpatrick J. Radiation dose-volume effects of optic nerves and chiasm. Int J Radiat Oncol Biol Phys. 2010 Mar 1;76(3 Suppl):S28–35. PMID: 20171514

35. Mayo C, Yorke E, Merchant TE. Radiation associated brainstem injury. Int J Radiat Oncol Biol Phys. 2010 Mar 1;76(3 Suppl):S36–41. PMCID: PMC2899702

